# Identification of genetic variants and their causal association underlying autism spectrum disorder

**DOI:** 10.1101/2024.09.27.24314463

**Authors:** Prashasti Yadav, Mona Srivastava, Parimal Das

## Abstract

**Background:** Autism spectrum disorder (ASD) is an early-onset neurodevelopmental disorder with a complex genetic architecture characterized by persistent deficits in social communication and social interaction along with restricted and repetitive behavior patterns, interests and activities. Over the years its prevalence has increased and affects 1% children globally. Etiology of ASD is remained unclear, but it is well recognized as a complex disorder involving interactions of several genes and environmental risk factors. Studies signify the role of whole exome sequencing in identifying rare genetic variation related to neurodevelopmental disorders and have greatly improved the chance of identifying known as well as novel responsible genes. This study aimed to identify genetic variant in a patient affected with autism spectrum disorder.

**Methods:** A male patient of age range 0-5 years was investigated and identified according to DSM-5 criteria for autism spectrum disorder. Peripheral blood sample was collected followed by genomic DNA extraction and whole exome sequencing. The functional impact of variant, gene interaction network and pathway enrichment analysis were done using *in silico* tools and databases.

**Results:** Two heterozygous missense variants one in *SHANK2* (c.401C>G) another in *CSNK2A1 (*c.572G>A) were identified. Both of these variants were predicted to be deleterious by pathogenicity prediction tool. Gene interaction network and pathway enrichment analysis revealed the involvement of *SHANK2* and *CSNK2A1* in various biological pathways such as protein binging, synaptogenesis, chromosome condensation and cell cycle regulation.

**Conclusion:** This study revealed a novel *SHANK2* variant along with a known *CSNK2A1* variant that expands the spectrum of genetic variants related to autism spectrum disorder and further enhances the understanding of the molecular mechanism underlying *SHANK2* and *CSNK2A1* pathophysiology.

## Introduction

Autism Spectrum Disorder (ASD) is a childhood neurodevelopment disorder characterized by persistence deficits in social communication and social interaction as well as restricted and repetitive patterns of behavior. It is often present with cognitive impairments, language development delay and a spectrum of comorbidities, including hyperactivity and anxiety disorder [1]. Over the past few years, its prevalence has increased and affects about one in 100 children globally [2]. It is more common in male children than female children [3]. Because of the absence of reliable biomarkers, diagnosis is most often based on behavioural assessment of the children and also the therapeutic options are predominantly restricted to behavioural interventions [4, 5]. Aetiology of ASD remains unclear but now it is well recognised as a complex disorder involving interactions of several genes and environmental risk factors [6,7,8]. Family and twin studies indicated the significant genetic basis susceptibility and heritability of ASD [9,10]. There is increasing evidence that the genes involved in pathogenesis of ASD converge to common biological pathways that include chromatin modifiers, translational regulators, synapse formation and cortical development [11].

Mutations in genes that encode synaptic cell adhesion molecules and scaffolding proteins, such as neuroligins (NLGN), neurexins (NRXN) and SHANK family protein, have been reported in patients with ASD [12]. These proteins play significance role in the synapse formation and stabilization [13]. *De novo* deleterious *SHANK2* and *SHANK3* variants have been identified in individuals with intellectual disability (ID), autism spectrum disorder (ASD), developmental delay, and attention deficit and hyperactivity disorder [14, 15,16].

*CSNK2A1* is also categorized as autism risk gene that is reported in the SFARI gene database as high confidence autism risk gene [17]. Rare *de novo CSNK2A1* variants have been identified in ASD probands from simplex families from the Simons Simplex Collection [18, 19] and also reported in children with an intellectual disability and dysmorphic facial features syndrome [20].

In present study two missense variants one in *SHANK2* (c.401C>G) and another in *CSNK2A1* (c.572G>A) were identified in a proband from a single Indian family by whole exome sequencing. In addition, we further explored the functional impact of variants and pathways involved to understand the molecular mechanisms of *SHANK2* and *CSNK2A1*. The findings, provide valuable insights for pathogenicity of genetic variants and shed light on the underlying molecular process involved in ASD and also confirm the efficacy of whole exome sequencing in detecting genetic variants in neurodevelopmental disorders.

## Material and Methods

### Recruitment of patient and sample collection

Patient was enrolled from eastern Uttar Pradesh, India. The peripheral blood sample was collected after obtaining the written informed consent from parents, as the proband was minor. The study protocol was approved by the Institutional Ethics Committee of Centre for Genetic Disorders, Institute of Science, Banaras Hindu University, Varanasi.

The diagnosis of ASD was done according to the American Psychiatric Association’s Diagnostic and Statistical Manual of Mental Disorders (DSM-5) criteria [1], and also evaluation was done using standard scale, IASQ (The Indian Autism Screening Questionnaire) [21].

### DNA Extraction, whole exome sequencing and variant filtration

Genomic DNA was extracted using standard salting out method. Whole exome sequencing was performed using Illumina next-generation sequencing. Sequencing of approximately 30Mb of the human exome, targeting approximately 99% of regions in CCDS and RefSeq was performed. Complete mitochondrial genome sequencing was also performed at a mean depth of 80-100X. Percentage of bases covered was at 20X depth >90% in the target region and for the mitochondrial genome at 1000-2000X depth. Variant was identified using GATK best practice framework. Duplicate reads were identified and removed, base quality was recalibrated and re-alignment were done using DRAGEN BIO IT platform. Variant annotation was done using published databases like OMIM, GWAS, GNOMAD, 1000Genome and ClinVar.

## *In silico* function prediction

Pathogenicity and functional impact of variants were determined using *in silico* tool, Mutation Taster [22].

### Interaction Network and Gene Set Enrichment Analysis

We used freely accessible and user-friendly web tools GeneMANIA (http://www.genemania.org/) and STRING (https://string-db.org/) for networks construction and analysis.

Gene Multiple Association Network Integration Algorithm (GeneMANIA) is a web-based tool for prediction of gene function based on single gene or gene set query [23, 24].

Search Tool for the Retrieval of Interacting Genes (STRING) is an online protein-protein interaction database curated from literature and predicted associations. [25].

Gene set enrichment analysis was done using Enrichr tool, it is a freely available comprehensive resource and a search engine for gene set [26].

## Results

### Clinical Description of Patient

This study includes a single Indian family. Proband is a male of 0-5 years age range, second child born out of non-consanguineous marriage. He has no family history of ASD or neurodevelopmental disorder. Clinical features include loss of eye contact, repetitive behaviour, delayed walking and facial dysmorphism. Brain MRI investigation was normal.

### WES analysis

We identified two heterozygous missense variants one in *SHANK2* (c.401C>G) and another in *CSNK2A1* (c.572G>A). The pathogenicity of both variants was predicted as deleterious by *in silico* tool, Mutation Taster [Table1]

**Table 1.** Variants identified in *SHANK2* and *CSNK2A1* genes in proband in this study.

| Gene & transcript | Location | Nucleotide Substitution | Amino Acid Change | Zygosity | Mutation taster | ClinVar | Population Frequency (ExAC/gnomAD) |
| --- | --- | --- | --- | --- | --- | --- | --- |
| *SHANK2* NM_012309. | Exon 4 | c.401C>G | p.Pro134Arg | Heterozygous | Disease causing | NA | 0/0 |
| 5 |  |  |  |  |  |  |  |
| *CSNK2A1*<br>NM_177559.<br>3 | Exon 9 | c.572G>A | Arg191Gln | Heterozygous | Disease causing | NA | 0/0 |

NA: Not available

### Interaction Network and Gene Set Enrichment Analysis

In order to understand the functional implication of *SHANK2* and *CSNK2A1* we performed gene interaction network and gene set enrichment analysis.

Gene network derived from GeneMANIA database using *SHANK2* and *CSNK2A1* as input genes, moreover another 20 strongly connected genes in network are, *CSNK2B, CSNK2A2, TOP2A, DVL1, PIN1, APC, DNM2, SSTR2, STX4, ARHGEF7, NCAPD2, NCAPG, NCAPH, SUPT16H, L1CAM, MAP1B, BAIAP2L1, MAPK3, CALD1* and *STK11*. [Figure 1 A]. STRING Protein-protein interaction (PPI) network analysis show the functional association among above proteins and revealed the involvement of these proteins in various pathways such as nervous system development, protein binding, chromosome condensation, and cell cycle regulation [Figure 1 B].

**Figure 1.**
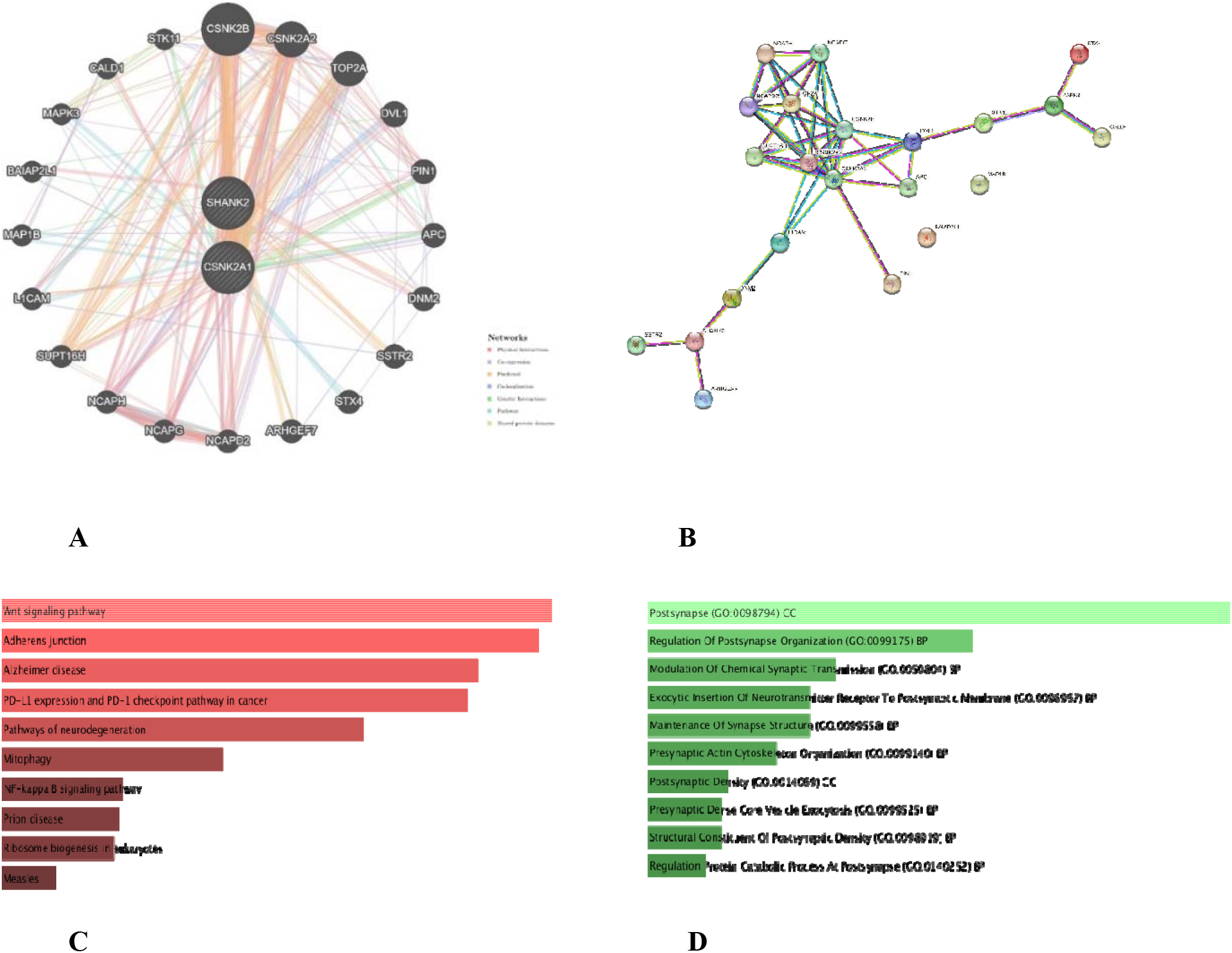
GeneMANIA gene network showing the top 20 strongly connected genes with *SHANK2* and *CSNK2A1*. **(A)** SRTING PPI network showing relationship for proteins (nodes) connected (with edges) according to their functional association. (**B)** Bar graph showing the top ten enriched KEGG pathways derived from Enrichr database. (**C)** Bar graph showing top ten Gene Ontologies (SynGO 2024, a database for Synaptic Gene Ontologies and Annotation) derived from Enrichr database. **(D)**

The top enriched KEGG pathways derived from Enrichr database for the above gene set are, Wnt signaling pathway *(CSNK2A1, APC, CSNK2A2, CSNK2B, DVL1)*, Adherens junction *(CSNK2A1, CSNK2A2, CSNK2B, MAPK3*), Alzheimer disease *(CSNK2A1, APC, CSNK2A2, CSNK2B, DVL1, MAPK3)*, PD-L1 expression and PD-1 checkpoint pathway in cancer *(CSNK2A1, CSNK2A2, CSNK2B, MAPK3)* and Pathways of neurodegeneration *(CSNK2A1, APC, CSNK2A2, CSNK2B, DVL1, MAPK3)* [Figure 1 C.]

The top enriched Synaptic Gene Ontologies derived from the Enrichr database are, Post synapse *(APC, CALD1, MAP1B, STX4)*, Regulation of post synapse organization *(DVL1, SHANK2)*, Modulation of chemical synaptic transmission *(L1CAM, MAPK3)*, Exocytic insertion of neurotransmitter receptor to postsynaptic membrane *(STX4)* and maintenance of synapse structure *(L1CAM)* [Figure 1 D].

## Discussion

Autism Spectrum Disorder (ASD) is a neurodevelopmental disorder with complex genetic architecture [8]. In the past few years, the development of genome sequencing technology has advanced the variant discovery. Whole exome sequencing has been used to identify rare and novel genetic variation related to neurodevelopmental disorder [27].

In present study we identified two missense variants one in *SHANK2 (*c.401C>G; p.Pro134Arg) another in *CSNK2A1* (c.572G>A; p.Arg191Gln) from the analysis of whole exome sequencing.

*SHANK2* (c.401C>G; p.Pro134Arg) is a heterozygous missense variantion. It is a novel variation, not observe in 1000 genomes, GnomeAD and ClinVar databases. *In silico* functional impact of this variant is predicted as deleterious.

*SHANK2* encodes a protein that is a member of SHANK family of synaptic protein essential for proper synapse formation, development and plasticity [28]. It is located in the postsynaptic density of glutamatergic synapses and encode scaffolding protein [29]. *SHANK* (SH3 and multiple ankyrin repeat domains protein) family genes comprising *SHANK1, SHANK2* and *SHANK3* have been linked to a spectrum of neurodevelopmental disorders [30].

*CSNK2A1* (c.572G>A, Arg191Gln) is a heterozygous missense variant. It is not observed in 1000 genomes, GnomeAD and ClinVar databases. *In silico* functional impact of this variant is predicted as deleterious. It is a known and previously reported variant [20] and also observed in SFARI gene database.

*CSNK2A1* encodes Casein kinase II subunit alpha, a constitutively active and acidophilic serine/threonine protein kinase complex, present in cells in a tetrameric form and composed of two catalytic subunits (α or its isoform α’) and two regulatory subunits (β) [31]. It is expressed in eukaryotic organism’s all tissues and is essential for normal embryonic development [32]. It phosphorylates large number of substrates and regulates numerous biological processes such as apoptosis, cell proliferation and the DNA damage response [33].

Rare *de novo CSNK2A1* variants have been identified in children with ASD, intellectual disability and dysmorphic facial features syndrome [18, 19, 20,].

Gene-network derived from GeneMANIA database show the top 20 strongly connected genes with *SHANK2* and *CSNK2A1* [Figure 1 A]. STRING PPI network analysis show the functional association among these proteins and revealed the involvement in various functions such as proteins binging, synaptogenesis, chromosome condensation and cell cycle regulation [Figure 1 B].

The gene set enrichment analysis using Enrichr tool [26] revealed the major pathways and Gene Ontologies for the above gene set. The top enriched KEGG pathways are, Wnt signaling pathway, Adherens junction, Alzheimer disease, expression and PD-1 checkpoint pathway in cancer and pathways of neurodegeneration [Figure 1 C]. The top enriched Gene Ontologies are Postsynapse organization, Modulation of chemical synaptic transmission, Exocytic insertion of neurotransmitter receptor to postsynaptic membrane and Maintenance of synapse structure [Figure 1 D].

## Conclusion

In summary, we identified a novel heterozygous missense *SHANK2* variant along with a known heterozygous missense *CSNK2A1* variant and utilized the existing bioinformatics tools and databases to gain insight into pathways related to autism spectrum disorder. Further, the functional characterization of identified variants in cell and animal models will be needed to understand the pathophysiology of ASD.

## Data Availability

All data produced in the present work are contained in the manuscript

